# Assessment of MYC Gene and WNT Pathway Alterations in Early-Onset Colorectal Cancer Among Hispanic/Latino Patients Using Integrated Multi-Omics Approaches

**DOI:** 10.1101/2024.12.05.24318588

**Authors:** FG Carranza, B Waldrup, Y Jin, Y Amzaleg, M Postel, DW Craig, JD Carpten, B Salhia, D Hernandez, N Gutierrez, CN Ricker, JO Culver, CE Chavez, MC Stern, L Baezconde-Garbanati, HJ Lenz, E Velazquez-Villarreal

**Affiliations:** City of Hope, Beckman Research Institute, Department of Integrative Translational Sciences, Duarte, CA; University of Southern California, Keck School of Medicine of USC, Department of Translational Genomics, Los Angeles, CA; City of Hope Comprehensive Cancer Center, Duarte, CA; University of Southern California, USC Norris Comprehensive Cancer Center, Los Angeles, CA; University of Southern California, Keck School of Medicine of USC, Division of Medical Oncology, Los Angeles, CA; Los Angeles General Medical Center, Los Angeles, CA; University of Southern California, Keck School of Medicine of USC, Department of Population and Public Health Sciences, Los Angeles, CA

## Abstract

Colorectal cancer (CRC) has increased at an alarming rate amongst younger (< 50 years) individuals. Such early-onset colorectal cancer (EOCRC) has been particularly notable within the Hispanic/Latino population. Yet, this population has not been sufficiently profiled in terms of two critical elements of CRC -- the MYC proto-oncogene and WNT signaling pathway. Here, we performed a comprehensive multi-omics analysis on 30 early-onset and 37 late-onset CRC (≥ 50 years) samples from Hispanic/Latino patients. Our analysis included DNA exome sequencing for somatic mutations, somatic copy number alterations, and global and local genetic similarity. Using RNA sequencing, we also assessed differential gene expression, cellular pathways, and gene fusions. We then compared our findings from early-onset Hispanic/Latino patient samples with publicly available data from Non-Hispanic White cohorts. Across all early-onset patients, which had a median 1000 Genomes Project Peruvian-in-Lima-like (1KG-PEL-like) genetic similarity proportion of 60%, we identified 41 WNT pathway genes with significant mutations. Six important examples were APC, TCF7L2, DKK1, DKK2, FZD10, and LRP5. Notably, patients with mutations in DKK1 and DKK2 had the highest 1KG-PEL-like proportion (79%). When we compared the Hispanic/Latino cohort to the Non-Hispanic White cohorts, four of these key genes -- DKK1, DKK2, FZD10, and LRP5 -- were significant in both risk association analyses and differential gene expression. Interestingly, early-onset tumors (vs. late-onset) exhibited distinct somatic copy number alterations and gene expression profiles; the differences included MYC and drug-targetable WNT pathway genes. We also identified a novel WNT gene fusion, RSPO3, in early-onset tumors; it was associated with enhanced WNT signaling. This integrative analysis underscores the distinct molecular features of EOCRC cancer in the Hispanic/Latino population; reveals potential avenues for tailored precision medicine therapies; and emphasizes the importance of multi-omics approaches in studying colorectal carcinogenesis. We expect this data to help contribute towards reducing cancer health disparities.

**Significance:** This study offers multi-omics profiling analysis of early-onset colorectal cancer (EOCRC) in an underserved community, explores the implications of *MYC* gene and WNT pathway alterations, and provides critical insights into cancer health disparities.

**ABSTRACT(short version):** Colorectal cancer (CRC) has risen at an alarming rate in early-onset (<50 yrs) cases. Both its increased incidence and higher rates of mortality are especially pronounced within the Hispanic/Latino population. Although the *MYC* gene and WNT signaling pathway are well-established in CRC, we lack sufficient data about what role these elements play in young Hispanic/Latino patients. Here, we assess how both the *MYC* gene and WNT pathway are altered in Hispanic/Latino patients with early-onset CRC. We analyzed 30 early-onset and 37 late-onset CRC samples using multi-omics approaches, including DNA exome and RNA sequencing. The strategy allowed us to identify significant differences between early and late-onset tumors. Specifically, early-onset CRC had prevalent alterations in WNT pathway genes*: APC, TCF7L2, DKK1, DKK2*, and *FZD10*. Unique mutational profiles were linked to a high proportion of Peruvians-from-Lima-like (1KG-PEL-like) genetic similarity. These findings highlight the need for targeted precision medicine approaches to address the distinct molecular characteristics of early-onset CRC in underrepresented populations.

## INTRODUCTION

Colorectal cancer (CRC) is especially difficult to treat when diagnosed before the age of 50. Termed early-onset CRC (EOCRC), it is usually aggressive and has poor prognoses (Siegel et al., 2023; Hofseth et al., 2020). As the incidence of EOCRC continues to rise at an alarming pace, studies have suggested that it harbors unique molecular characteristics vs. late-onset CRC (≥ 50 years; LOCRC) (Sinicrope et al., 2022; Pearlman et al., 2017; REACCT collaborative et al., 2021; Mork et al., 2015; Cercek et al., 2021; Lieu et al., 2019). EOCRC (vs. LOCRC) more frequently exhibits chromosomal deletions and copy number variations (Berg et al., 2010; Brea-Fernandez et al., 2017). Significant progress thus has been made in understanding EOCRC across all CRC patients. Yet, a subset of these patients -- the Hispanic/Latino population -- remains understudied. This demographic appears to harbor unique molecular characteristics relevant to disease outcomes (Rhead et al., 2024; Pérez-Mayoral et al., 2019; Kato et al., 2002; Mauri et al., 2019). Understanding the genetic underpinnings of CRC in Hispanic/Latino patients is crucial for addressing cancer disparities and improving targeted treatment strategies (Monge et al., 2024; Carranza FG et al., 2024)

The pathogenesis of EOCRC is critically affected by both the MYC gene and WNT signaling pathway. MYC is a key oncogenic target of the WNT/β-catenin pathway and is frequently deregulated in CRC (Rennoll et al., 2015; Han et al., 2019). Over the years, MYC has been more frequently detected in CRC with somatic copy number alterations (SCNAs). A particularly high prevalence of SCNAs in MYC has been seen in younger CRC patients (You et al., 2020; Al-Kuraya et al., 2007; Marx et al., 2022; Pan et al., 2020). Furthermore, EOCRC has exhibited alterations in MYC regulatory genes. Two examples are mutations in 1) an E3 ubiquitin ligase called MYCBP2, which may be involved in regulating MYC transcription (Tricoli et al., 2018; Guo et al., 1998); 2) a tumor suppressor ubiquitin ligase called FBXW7, which mediates the degradation of MYC and other oncoproteins (Kothari et al., 2016; Tricoli et al., 2018). Despite these findings, additional research is needed to fully understand the functional implications of these mutations (Yeh et al., 2018). Collectively, these insights suggest a possible WNT-independent increase in MYC activity in EOCRC, which may contribute to the distinct biology of the disease in younger Hispanic/Latino patients.

Here, we mapped the molecular landscape of EOCRC in Hispanic/Latino patients using whole-exome sequencing (WES) and RNA-sequencing approaches. For this demographic, we compared how the molecular features differed when CRC was early onset vs. late onset. We then compared whether EOCRC in Hispanic/Latino patients had different molecular features than EOCRC in non-Hispanic White patients. This comparative analysis allowed us to identify unique genetic and molecular alterations. Ultimately, we expect these results can lead to new strategies for guiding precision medicine and improving outcomes for Hispanic/Latino CRC patients.

## RESULTS

### Patient demographics and tumor characteristics

In this study cohort for the EOCRC Moonshot project included 30 early- and 37 late-onset Hispanic/Latino patients (**Table 1**). The early-onset group had approximately twice as many male than female patients. The late-onset group had a majority of female patients. Most tumors were microsatellite stable (MSS): 73% of early- and 86.5% of late-onset groups. Fewer tumors were microsatellite instable (MSI): 26.7% of early- and 13.5% of late-onset groups. Few tumors were hypermutated: 36.7% of early-onset and 13.5% of late-onset groups. Tumor localization was primarily in the colon for both groups, with 63.3% of early- and 62.2% of late-onset tumors occurring there. According to pathological staging, 33.3% of early- and 24.3% of late-onset tumors were either in Stage I or II. In contrast, 43.3% of early- and 43.2% of late-onset tumors were either in Stage III or IV.

**Table 1.**
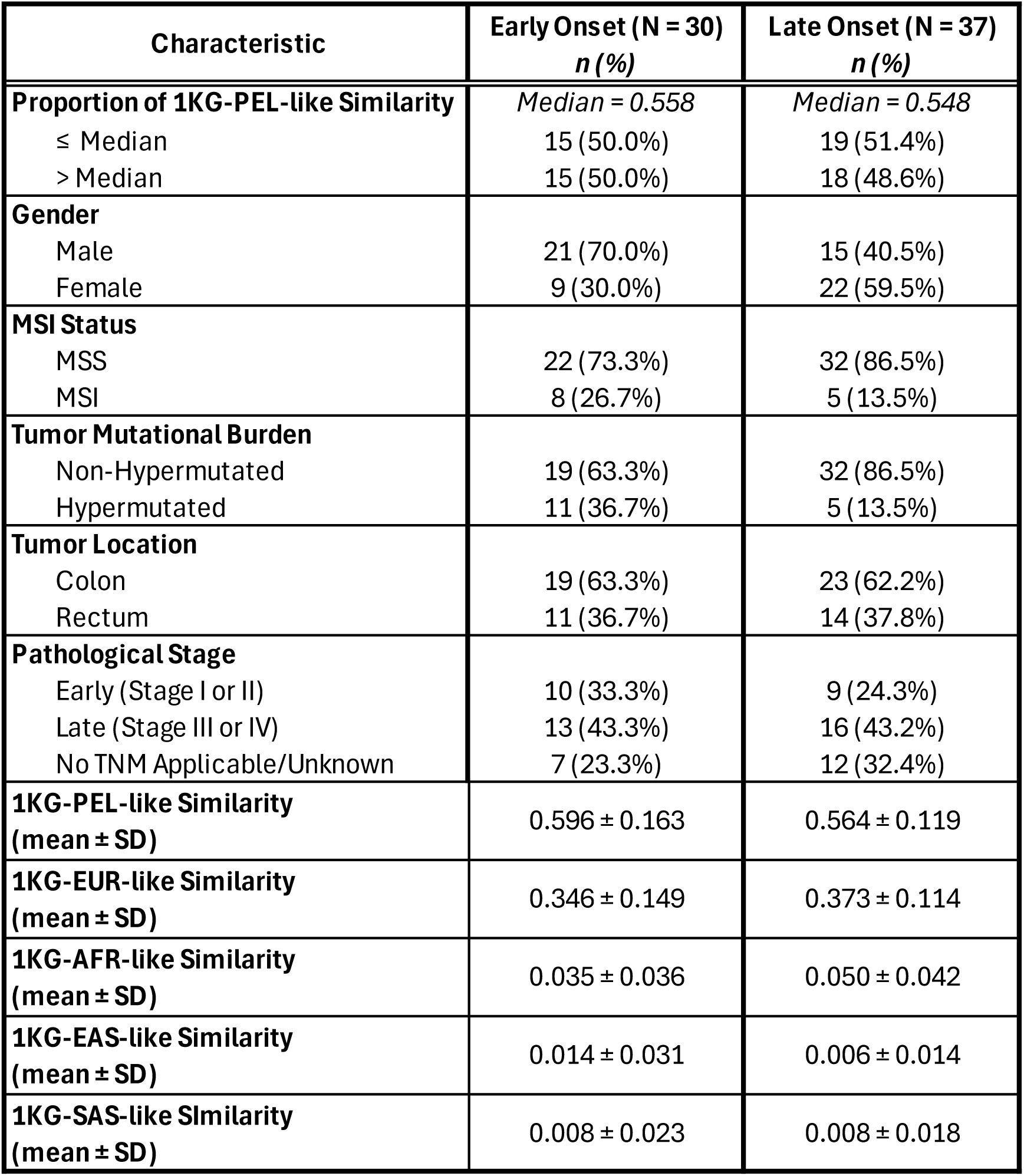
Patient Demographics, Clinical Characteristics and Genomic Similarity Proportions. This table provides details on patient demographics and clinical characteristics, along with the proportions of genetic similarity, classified into five super populations: 1KG-PEL-like Similarity, 1KG-EUR-like Similarity, 1KG-AFR-like Similarity, 1KG-EAS-like Similarity, and 1KG-SAS-like Similarity. The admixed genomic similarity proportions offer insights into the continental ancestral origins of the patients’ genomes.

| Characteristic | Early Onset (N = 30)<br>*n* (%) | Late Onset (N = 37)<br>*n* (%) |
| --- | --- | --- |
| **Proportion of 1KG-PEL-like Similarity** | *Median = 0.558* | *Median = 0.548* |
| ≤ Median | 15 (50.0%) | 19 (51.4%) |
| > Median | 15 (50.0%) | 18 (48.6%) |
| **Gender** |  |  |
| Male | 21 (70.0%) | 15 (40.5%) |
| Female | 9 (30.0%) | 22 (59.5%) |
| **MSI Status** |  |  |
| MSS | 22 (73.3%) | 32 (86.5%) |
| MSI | 8 (26.7%) | 5 (13.5%) |
| **Tumor Mutational Burden** |  |  |
| Non-Hypermutated | 19 (63.3%) | 32 (86.5%) |
| Hypermutated | 11 (36.7%) | 5 (13.5%) |
| **Tumor Location** |  |  |
| Colon | 19 (63.3%) | 23 (62.2%) |
| Rectum | 11 (36.7%) | 14 (37.8%) |
| **Pathological Stage** |  |  |
| Early (Stage I or II) | 10 (33.3%) | 9 (24.3%) |
| Late (Stage III or IV) | 13 (43.3%) | 16 (43.2%) |
| No TNM Applicable/Unknown | 7 (23.3%) | 12 (32.4%) |
| **1KG-PEL-like Similarity**<br>**(mean ± SD)** | 0.596 ± 0.163 | 0.564 ± 0.119 |
| **1KG-EUR-like Similarity**<br>**(mean ± SD)** | 0.346 ± 0.149 | 0.373 ± 0.114 |
| **1KG-AFR-like Similarity**<br>**(mean ± SD)** | 0.035 ± 0.036 | 0.050 ± 0.042 |
| **1KG-EAS-like Similarity**<br>**(mean ± SD)** | 0.014 ± 0.031 | 0.006 ± 0.014 |
| **1KG-SAS-like Similarity**<br>**(mean ± SD)** | 0.008 ± 0.023 | 0.008 ± 0.018 |

### Genomic analysis

We assessed CRC in Hispanic/Latino patients in terms of genetic similarity, mutational landscape, and chromosomal alterations. Specifically, we conducted comprehensive bioinformatics analyses on whole exome sequencing (WES) data from 30 early-onset and 37 late-onset Hispanic/Latino tumor-normal pairs. Both tumor and matched normal tissues were sequenced using the same platform. This approach was used to ensure the data was consistent. Detailed quality metrics and sequencing statistics are presented in **Supplementary Table 1**. The results provide a robust foundation for subsequent genomic analyses.

### Genetic similarity analysis

Both early-onset and late-onset CRC patients self-reported race and ethnicity. We also complemented this data with a genetic similarity analysis to estimate genetic similarity proportions and admixture. genetic similarity proportions were determined for each group via germline genetic similarity Informative Markers from whole-exome sequencing (WES) data (**Fig. 1a**). The average genetic similarity composition for early-onset CRC cases was as follows: 60% 1000 Genomes Project Peruvian-in-Lima-like (1KG-PEL-like), 35% 1000 Genomes Project European-like (1KG-EUR-like), 4% 1000 Genomes Project African-like (1KG-AFR-like), 1% 1000 Genomes Project South-Asian-like (1KG-SAS-like), and 1% 1000 Genomes Project East-Asian-like (1KG-EAS-like). For late-onset CRC cases, the proportions were 56% 1KG-PEL-like, 37% EUR, 5% AFR, and 1% SAS, and 1% EAS (**Table 1**).

**Figure 1:**
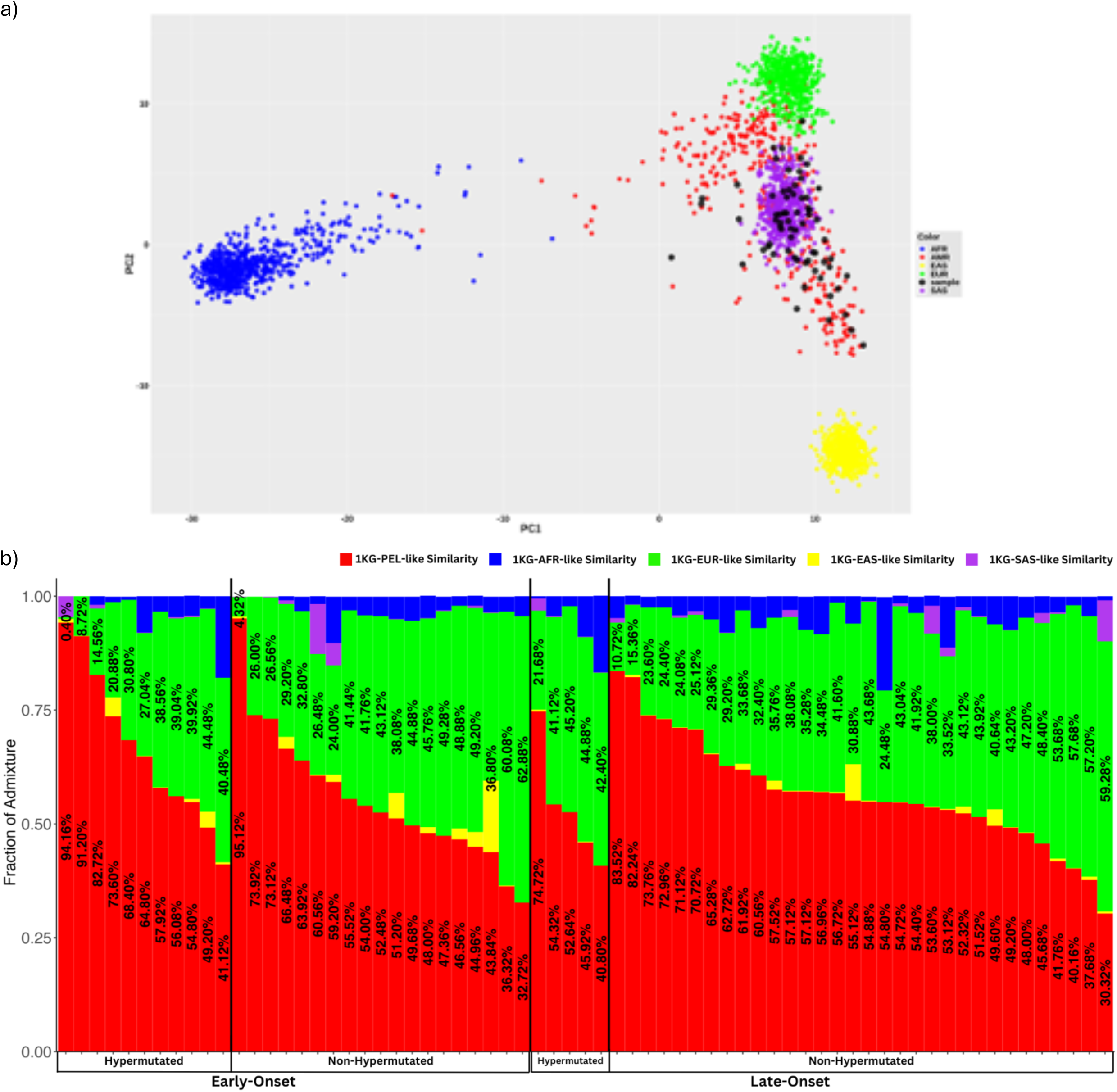
Global Ancestry and Chromosome-Level Admixture Patterns. **a)** Global ancestry principal component analysis (PCA) plot. This PCA plot illustrates the global ancestry of our Hispanic/Latino colorectal cancer cohort of 67 individuals, represented by black points, in the context of five super populations: African (1KG-AFR-like, blue), Peruvian-Lima (1KG-PEL-like, red), East Asian (1KG-EAS-like, yellow), European (1KG-EUR-like, green), and South Asian (1KG-SAS-like, purple). The plot displays the distribution of these samples along two principal components, PC1 and PC2, which captures the majority of variance in the genetic data. This visualization helps to contextualize the ancestry composition of our cohort relative to these major global populations. **b)** This figure illustrates the similarity frequencies in each tumor sample from a cohort of 67 Hispanic/Latino (H/L) colorectal cancer (CRC) patients. The genomes are categorized into five super populations, same as section a. Samples are stratified by age of diagnosis into two groups, early-onset and late-onset, with the proportion of 1KG-PEL-like and 1KG-EUR-like ancestry included.

In both early-onset and late-onset groups, all individuals self-identified as Hispanic/Latino and exhibited genetic similarity consistent with this identification. A high percentage of hypermutated tumors (both early and late) had an 1KG-PEL-like> 50% (**Fig. 1b**). In contrast, a much lower percentage of non-hypermutated early-onset tumors had an 1KG-PEL-like> 55.8%; and most non-hypermutated late-onset tumors had a 1KG-PEL-like ≤ 54.8% (**Fig. 1b**). To complement this approach, which estimates genetic similarity proportions across the entire genome, we conducted a Local genetic similarity Analysis. This approach allowed us to 1) determine genetic similarity proportions at the chromosomal level and 2) identify specific WNT pathway genes associated with higher proportions of 1KG-PEL-like. In early-onset tumors, genes such as RNF43, MYCBPAP, WNT9B, WNT3, AXIN2, and SOX9 exhibited elevated proportions of 1KG-PEL-like in both chromosomes. In contrast, late-onset tumors displayed a distinct pattern, with frequent elevations in genes like TCF7L2, GSK3B, DKK1, WNT8B, WNT9B, MYCBPAP, FZD8, RNF43, WNT3, SOX9, FZD6, MYC, and AXIN2 (**Supplementary Fig. 1**). The comparison reveals that genes such as RNF43, WNT9B, WNT3, AXIN2, and SOX9 are consistently altered in both early-onset and late-onset tumors. However, the late-onset group shows additional alterations in genes like TCF7L2, GSK3B, DKK1, WNT8B, FZD8, FZD6, and MYC. These distinctions highlight potential differences in WNT signaling for early and late-onset tumors in Hispanic/Latino patients. The results suggest that these variations may contribute to the unique biological characteristics observed in these subgroups.

We then explored clinical and molecular differences between early-onset and late-onset Hispanic/Latino groups. We focused on tumor characteristics including location, stage, microsatellite status, tumor mutational burden, and mutations in MYC and WNT pathway genes (**Supplementary Table 2**). Among these comparisons, only tumor mutational burden (TMB) showed a statistically significant difference between early-onset and late-onset groups. TMB was found to be approximately four times more prevalent in the late-onset group, though the precise magnitude of risk increase remains uncertain due to a wide confidence interval (**Supplementary Table 3**).

Finally, we assessed the impact of genetic similarity on the clinical and molecular characteristics of CRC for these patients. Specifically, we examined tumors from patients with varying proportions of 1KG-PEL-like and 1KG-EUR-like genetic similarity. We focused on key molecular markers such as MYC and WNT pathway genes (**Supplementary Table 4**). Tumor location showed a statistically significant difference when comparing early-onset samples with higher (>55.8%) and lower (≤55.8%) proportions of 1KG-PEL-like and late-onset samples with higher (>54.8%) and lower (≤54.8%) proportions of 1KG-PEL-like genetic similarity. Additionally, the FBXW7 mutation status exhibited significant variation within the late-onset group across different levels of 1KG-PEL-like genetic similarity (**Supplementary Table 4**). However, this variation did not reach statistical significance in the association analysis (**Supplementary Table 5**).

### Gene mutation analysis

To characterize the mutational landscape, we analyzed WES data from 30 early-onset and 37 late-onset CRC tumors along with matched normal tissue. The somatic mutation rates varied significantly among the samples. Notably, two early-onset samples exhibited mutation rates exceeding 100 mutations per 10^6 bases. Conversely, two late-onset samples had mutation rates below 1 per 10^6 bases. We categorized the scenarios into two groups. First, we termed cases as “non-hypermutated” when they had mutation rates ≤ 10 per 10^6 bases: 63% in early-onset and 86% in late-onset. Second, we termed cases as “hypermutated” when they had mutation rates > 10 per 10^6 bases (37% in early-onset and 14% in late-onset, **Fig. 2a**, **Table 1**). Among the hypermutated tumors, 73% in the early-onset group and 100% in the late-onset group exhibited MSI. Seventy five percent of hypermutated early-onset tumors had a 1KG-PEL-like > 55.8% versus 20% in late-onset cases (1KG-PEL-like > 54.8%) (**Fig. 2a**).

**Figure 2:**
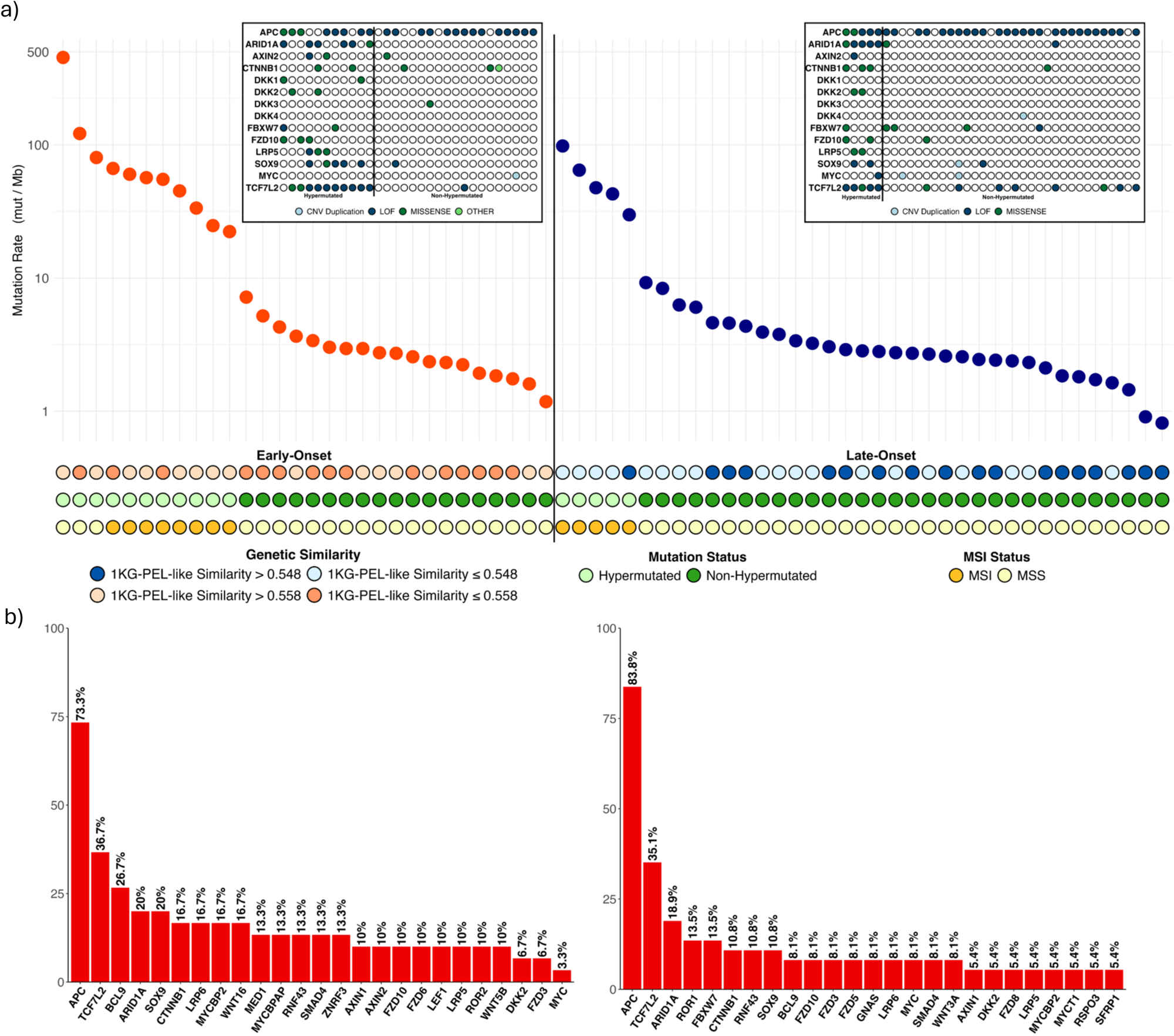
Mutation Frequencies in CRC Tumors from H/L Patients. **a)** This panel shows the mutation frequencies in each of the tumor samples from 67 Hispanic/Latino colorectal cancer patients. The samples are categorized as early-onset or late-onset. The color codes represent different attributes: dark orange for early-onset samples with an 1KG-PEL-like similarity proportion less than or equal to the median, light orange for early-onset samples with an 1KG-PEL-like similarity proportion greater than the median, light blue for late-onset samples with an 1KG-PEL-like similarity proportion less than or equal to the median, dark blue for late-onset samples with an 1KG-PEL-like similarity proportion greater than the median, light green for hypermutated, dark green for non-hypermutated, dark yellow for MSI, and light yellow for MSS. The inset highlights mutations in genes apart of the Wnt pathway among the early-onset and late-onset groups, with the sample order consistent with the main graph. **b)** This panel depicts the significantly mutated genes in early-onset and late-onset tumors.

We analyzed mutations in the MYC gene and WNT pathway genes as reported in the TCGA CRC project (Cancer Genome Atlas Network et al. 2012). In our early-onset cohort, hypermutated tumors exhibited the following mutation proportions: MYC (0.0%), APC (72.7%), ARID1A (54.5%), AXIN2 (18.2%), CTNNB1 (18.2%), DKK1 (18.2%), DKK2 (18.2%), DKK3 (0.0%), DKK4 (0.0%), FBXW7 (18.2%), FZD10 (27.3%), LRP5 (27.3%), SOX9 (45.5%), and TCF7L2 (90.9%). In contrast, the late-onset cohort with hypermutated MSI tumors showed different proportions: MYC (20.0%), APC (100.0%), ARID1A (100.0%), AXIN2 (20.0%), CTNNB1 (60.0%), DKK1 (0.0%), DKK2 (40.0%), DKK3 (0.0%), DKK4 (0.0%), FBXW7 (20.0%), FZD10 (40.0%), LRP5 (40.0%), SOX9 (40.0%), and TCF7L2 (100.0%). These findings align with previous reports and highlight notable differences between early-onset and late-onset CRC (Marx et al. 2023). Markedly, in our study, early-onset tumors with MYC aberrations had a lower average proportion of 1KG-PEL-like (36%) vs. late-onset tumors with MYC aberrations (55%). Furthermore, early-onset tumors with mutations in WNT pathway genes -- APC and TCF7L2 -- exhibited higher 1KG-PEL-like (60% and 63% on average, respectively) vs. the late-onset group (56% and 59% on average) (**Supplementary Table 6**).

In both early-onset and late-onset hypermutated and non-hypermutated colorectal cancers, we observed recurrent somatic mutations in MYC and WNT pathway genes. Our results are consistent with previous findings from the TCGA CRC project (Cancer Genome Atlas Network et al., 2012) (**Fig. 2b**). The most frequently mutated genes in early-onset tumors were APC, TCF7L2, BCL9, ARID1A, and SOX9. In late-onset tumors, the top mutated genes included APC, TCF7L2, ARID1A, ROR1, and FBXW7. These genes are crucial for various mechanisms that are directly relevant to treating CRC **[20]**. Notably, the mutation rates of the MYC gene did not significantly differ between early-onset and late-onset groups (8% versus 3%, P=0.76). However, the DKK1 gene was significantly more frequently mutated in early-onset tumors compared to late-onset tumors (18.2% versus 0%, P=0.01). Conversely, three genes were more frequently mutated in late-onset tumors than in early-onset tumors. More frequent mutations in 1) APC (72.7% versus 100%, p=0.0003) suggest that alterations in the WNT signaling pathway may be more critical in the pathogenesis of CRC in older patients; 2) ARID1A (54.5% versus 100%, p=1.486×10⁻⁶) implicate epigenetic changes in late CRC tumorigenesis; 3) CTNNB1 (18.2% versus 60%, p=0.0005), emphasizes the importance of WNT signaling for late CRC because CTNNB1 encodes the crucial signaling component β-catenin. Altogether, the mutation frequency of key MYC and WNT pathway genes are significantly different in early-onset vs. late-onset CRC in Hispanic/Latino populations (**Fig. 2b**).

### Comparison with early-onset CRC cohorts from public databases

We conducted a comparative analysis of mutation frequencies in MYC and WNT pathway genes for disparate populations: we compared our cohort of Hispanic/Latino patients, who have EOCRC, to six publicly available, CRC genomic datasets. These datasets are predominantly comprised of Non-Hispanic White patients; they include 66 cases from MSK-NatCommun, 851 cases from MSK-JNCI, 10 cases from DFCI-CellReports, 77 cases from TCGA-PanCancerAtlas, 122 cases from MSK-Gastroenterology, and 17 cases from MSK-JCO-PrecisOncol (**Table 2**). Across all seven datasets, including data from our cohort, the three most frequently mutated genes, after APC, were ARID1A, CTNNB1, and TCF7L2; significant differences were observed in four of the databases. Four other features were notable in our analysis of gene mutation frequencies. 1) Three of the databases exhibited distinct FZD10 and LRP5; these two genes were frequently mutated in our Hispanic/Latino cohort. 2) Two databases had notable differences in AXIN2, DKK1, DKK2, and SOX9. 3) One dataset showed significant differences in APC, DKK3, DKK4, and FBXW7 mutation frequency. 4) The mutation frequency of the tumor initiation gene APC in our early-onset cohort was 73.33%. This is significantly higher than the 19.7% observed in the MSK-NatCommun cohort, but did not differ from the other cohorts. These findings suggest potential ethnic-specific genetic variations in early-onset CRC.

**Table 2.** Mutation frequencies across CRC studies. This table includes gene names, mutation frequencies from the current Moonshot study (N=30), and frequencies from external studies: MSK-NatCommun (N= 66), MSK-JNCI (N= 851), DFCI-CellReports (N= 10), TCGA-PanCancerAtlas (N= 77), MSK-Gastroenterology (N= 122), MSK-JCO-PrecisOncol (N= 17). The included p-values demonstrate the significance levels when comparing frequencies with our study.

|  | Moonshot Study (N = 30) | MSK-NatCommun (N = 66) |  | MSK-JNCI (N = 851) |  | DFCI-CellReports (N = 10) |  |
| --- | --- | --- | --- | --- | --- | --- | --- |
| Gene | Mutated *n* (%) | Mutated *n* (%) | p-value | Mutated *n* (%) | p-value | Mutated *n* (%) | p-value |
| APC | 22 (73.33%) | 13 (19.70%) | **0.000001351** | 648 (76.15%) | 0.891 | 7 (70.00%) | 1 |
| ARID1A | 6 (20.00%) | 3 (4.55%) | **0.02463** | 66 (7.76%) | **0.03873** | 0 (0.00%) | 0.3074 |
| AXIN2 | 3 (10.00%) | 0 (0.00%) | **0.02842** | 38 (4.47%) | 0.159 | 0 (0.00%) | 0.5597 |
| CTNNB1 | 5 (16.67%) | 0 (0.00%) | **0.002331** | 52 (6.11%) | **0.05330** | 0 (0.00%) | 0.3059 |
| DKK1 | 2 (6.67%) | 0 (0.00%) | 0.09539 | 0 (0.00%) | **0.001122** | 0 (0.00%) | 1 |
| DKK2 | 2 (6.67%) | 0 (0.00%) | 0.09539 | 0 (0.00%) | **0.001122** | 0 (0.00%) | 1 |
| DKK3 | 1 (3.33%) | 0 (0.00%) | 0.3125 | 0 (0.00%) | **0.03405** | 0 (0.00%) | 1 |
| DKK4 | 1 (3.33%) | 0 (0.00%) | 0.3125 | 0 (0.00%) | **0.03405** | 0 (0.00%) | 1 |
| FBXW7 | 2 (6.67%) | 10 (15.15%) | 0.330 | 109 (12.81%) | 0.413 | 3 (30.00%) | 0.08929 |
| FZD10 | 3 (10.00%) | 0 (0.00%) | **0.02842** | 0 (0.00%) | **0.00003575** | 0 (0.00%) | 0.5597 |
| LRP5 | 3 (10.00%) | 0 (0.00%) | **0.02842** | 0 (0.00%) | **0.00003575** | 1 (10.00%) | 0.5597 |
| MYC | 1 (3.33%) | 1 (1.52%) | 0.530 | 6 (0.71%) | 0.216 | 0 (0.00%) | 1 |
| SOX9 | 6 (20.00%) | 3 (4.55%) | **0.02463** | 80 (9.40%) | 0.108 | 1 (10.00%) | 0.656 |
| TCF7L2 | 11 (36.67%) | 0 (0.00%) | **0.0000006194** | 121 (14.22%) | **0.001775** | 1 (10.00%) | 0.2307 |

|  | Moonshot Study (N = 30) | TCGA-PanCancerAtlas (N = 77) |  | MSK-Gastroenterology (N = 122) |  | MSK-JCO-PrecisOncol (N = 17) |  |
| --- | --- | --- | --- | --- | --- | --- | --- |
| Gene | Mutated *n* (%) | Mutated *n* (%) | p-value | Mutated *n* (%) | p-value | Mutated *n* (%) | p-value |
| APC | 22 (73.33%) | 52 (67.53%) | 0.988 | 80 (65.57%) | 0.807 | 15 (88.24%) | 0.282 |
| ARID1A | 6 (20.00%) | 9 (11.69%) | 0.422 | 7 (5.74%) | **0.033** | 6 (35.29%) | 0.420 |
| AXIN2 | 3 (10.00%) | 7 (9.09%) | 1 | 2 (1.64%) | **0.053** | 2 (11.76%) | 1 |
| CTNNB1 | 5 (16.67%) | 8 (10.39%) | 0.573 | 6 (4.92%) | **0.067** | 3 (17.65%) | 1 |
| DKK1 | 2 (6.67%) | 1 (1.30%) | 0.189 | 0 (0.00%) | **0.03791** | 0 (0.00%) | 0.5282 |
| DKK2 | 2 (6.67%) | 0 (0.00%) | 0.07671 | 0 (0.00%) | **0.03791** | 0 (0.00%) | 0.5282 |
| DKK3 | 1 (3.33%) | 1 (1.30%) | 0.484 | 0 (0.00%) | 0.1974 | 0 (0.00%) | 1 |
| DKK4 | 1 (3.33%) | 2 (2.60%) | 0.484 | 0 (0.00%) | 0.1974 | 0 (0.00%) | 1 |
| FBXW7 | 2 (6.67%) | 14 (18.18%) | 0.226 | 11 (9.02%) | 1 | 8 (47.06%) | **0.002187** |
| FZD10 | 3 (10.00%) | 6 (7.79%) | 0.708 | 0 (0.00%) | **0.007076** | 0 (0.00%) | 0.2923 |
| LRP5 | 3 (10.00%) | 2 (2.60%) | 0.133 | 0 (0.00%) | **0.007076** | 0 (0.00%) | 0.2923 |
| MYC | 1 (3.33%) | 3 (3.90%) | 1 | 1 (0.82%) | 0.357 | 1 (5.88%) | 1 |
| SOX9 | 6 (20.00%) | 9 (11.69%) | 0.422 | 7 (5.74%) | **0.033** | 4 (23.53%) | 1 |
| TCF7L2 | 11 (36.67%) | 9 (11.69%) | **0.0069** | 4 (3.28%) | **0.00000253** | 7 (41.18%) | 0.766 |

We further assessed somatic mutations in CRC cases using data from the AACR Project GENIE dataset (Table 3); GENIE includes self-reported race and ethnicity information. We combined cases from our early-onset cohort with those from both the early-onset TCGA-PanCancer and early-onset AACR GENIE datasets. This allowed us to stratify the data by self-identified race. The combined dataset comprised 64 patients (14%) who self-identified as Hispanic/Latino and 381 patients (86%) who self-identified as Non-Hispanic White (**Table 3**). Within this racially stratified cohort, we identified six statistically significant differences in the frequency of mutations of four key WNT pathway genes: DKK1, DKK2, FZD10, and LRP5 (**Table 3**).

**Table 3:** Mutation Frequencies in Hispanic/Latino and Non-Hispanic White Cases. This table compares mutation frequencies in Hispanic/Latino (H/L) and Non-Hispanic White (NHW) cases across three datasets: our Moonshot study, the AACR Project GENIE dataset, and the TCGA-PanCancer project. It lists gene names apart of the Wnt/β-catenin pathway along with their respective mutation frequencies as percentages for each racial group. The final column presents p-values, indicating the statistical significance of the observed differences in mutation frequencies between H/L and NHW patients.

| Gene | H/L Mutations<br>(N = 64) | NHW Mutations<br>(N = 381) | p-value |
| --- | --- | --- | --- |
| **APC** | 49 (76.56%) | 291 (76.38%) | 1 |
| **ARID1A** | 9 (14.06%) | 43 (11.29%) | 0.6676 |
| **AXIN2** | 6 (9.38%) | 16 (4.20%) | 0.1455 |
| **CTNNB1** | 8 (12.50%) | 25 (6.56%) | 0.1557 |
| **DKK1** | 2 (3.13%) | 1 (0.26%) | **0.0555** |
| **DKK2** | 2 (3.13%) | 0 (0.00%) | **0.02041** |
| **DKK3** | 1 (1.56%) | 1 (0.26%) | 0.2672 |
| **DKK4** | 1 (1.56%) | 1 (0.26%) | 0.2672 |
| **FBXW7** | 6 (9.38%) | 60 (15.75%) | 0.2530 |
| **FZD10** | 4 (6.25%) | 2 (0.52%) | **0.0047** |
| **LRP5** | 3 (4.69%) | 2 (0.52%) | **0.0230** |
| **MYC** | 2 (3.13%) | 11 (2.89%) | 1 |
| **SOX9** | 9 (14.06%) | 48 (12.60%) | 0.6901 |
| **TCF7L2** | 14 (21.88%) | 51 (13.39%) | 0.1123 |

### Somatic copy-number alterations analysis

We assessed both early-and late-onset colorectal tumors from Hispanic/Latino patients for somatic copy-number alterations (SCNAs) (**Fig. 3**)

**Figure 3:**
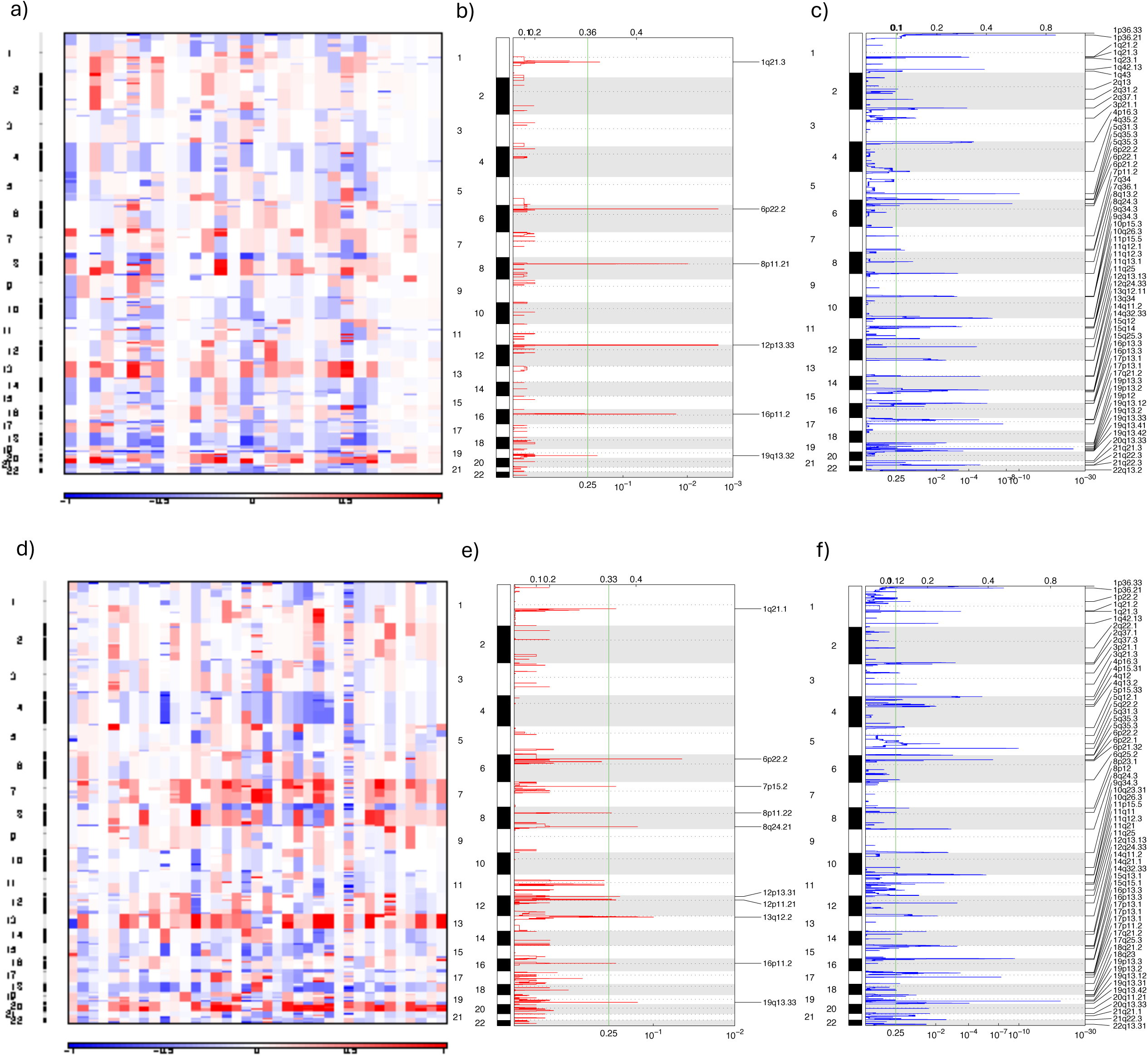
Somatic Copy Number Alterations (SCNAs) Analysis of Genomic Changes in 30 Early-Onset and 37 Late-Onset Hispanic/Latino Colorectal Cancer Patients. **a)** Heatmap of SCNAs: This panel displays SCNAs at both chromosomal and sub-chromosomal levels across 30 early-onset tumor samples from Hispanic/Latino colorectal cancer patients. Deletions (losses) are indicated in blue, while insertions (gains) are shown in red. **b)** Focal Deletions: This figure highlights the focal deletions identified across 30 early-onset tumor samples. **c)** Focal Amplifications: This figure presents the focal amplifications observed in the same set of tumor samples. **d)** Heatmap of SCNAs: This panel displays SCNAs at both chromosomal and sub-chromosomal levels across 37 late-onset tumor samples from Hispanic/Latino colorectal cancer patients. Deletions (losses) are indicated in blue, while insertions (gains) are shown in red. **e)** Focal Deletions: This figure highlights the focal deletions identified across 37 late-onset tumor samples. **f)** Focal Amplifications: This figure presents the focal amplifications observed in the same set of tumor samples.

In our analysis, some SCNAs were specific to late-onset tumors; some were observed for both groups; and others were unique to the early-onset tumors. We describe these findings in that order. 1) Late-onset tumors (vs. early-) had significantly more SCNAs (**Fig. 3a vs. Fig. 3d**). Moreover, the late-onset tumors alone -- and not the early-onset -- exhibited a deletion of the MYC gene at the chromosomal locus 8q24.21 (**Fig. 3e**). The late-onset tumors also displayed a broader spectrum of deletions across the following genes: FZD1 (7q21.2) and WNT2 (7q31.2) on chromosome 7; DKK1 (10q11.2) and FZD8 (10p11.22) on chromosome 10; and WNT1 (12q13.12) and FZD10 (12q24.33) on chromosome 12. These were observed with corresponding amplifications. 2) Both patient groups exhibit significant deletions and amplifications in critical genes such as CTNNB1 (3p22.1), WNT7A (3p25.1) on chromosome 3, and APC (5q22.2) on chromosome 5. 3) Only early-onset CRC exhibited changes in several WNT pathway genes. These pathways may be involved in driving tumorigenesis in the younger demographic. Early-onset CRC tumors also showed prominent deletions in genes like DKK4 (8p11.21) and FZD6 (8q22.3) on chromosome 8; and TCF7L2 (10q25.2) on chromosome 10. As before, these SCNAs were observed with corresponding amplifications, indicating their dual role in cancer development. Some of these genes play crucial roles in the WNT signaling pathway: particularly DKK1, the Frizzled receptors (i.e. FZD) genes, and TCF7L2. These are being investigated for therapeutic targeting.

Altogether, our findings suggest both early-onset and late-onset CRC may involve similar WNT pathway genes. However, the specific patterns of alteration and their biological roles differ between the two age groups. Our results suggest distinct mechanisms drive tumorigenesis in early versus late-onset CRC. This has important implications for age-specific therapeutic strategies.

### Transcriptomics analysis

We conducted RNA sequencing analysis on 30 early-onset colorectal tumors from Hispanic/Latino patients. We compared our results to data from 30 early-onset tumors from Non-Hispanic White patients and 37 late-onset tumors from the Cancer Moonshot project. For these diverse groups, we aimed to define two features: 1) gene expression profiles; 2) fusions involving both the MYC gene and WNT pathway genes.

### Comparison of Early-Onset and Late-Onset CRC Tumors in Hispanic/Latino patients

For this comparison, we employed a differential gene expression analysis (**Supplementary Fig. 2**). Among the WNT pathway genes, DKK1 emerged as the only statistically significant gene, with a p-value of <0.05 (**Fig. 4a, b**). DKK1, a known tumor suppressor gene, was found to be downregulated in early-onset tumors. Thus, in early-onset tumors, DKK1 may be less capable of inhibiting cell proliferation and tumor growth. Notably, the downregulation of DKK1 has been linked to poor prognosis in various cancers, including colorectal cancer. This highlights its potential impact on the aggressive nature of early-onset CRC (Gurney et al. 2012).

**Figure 4:**
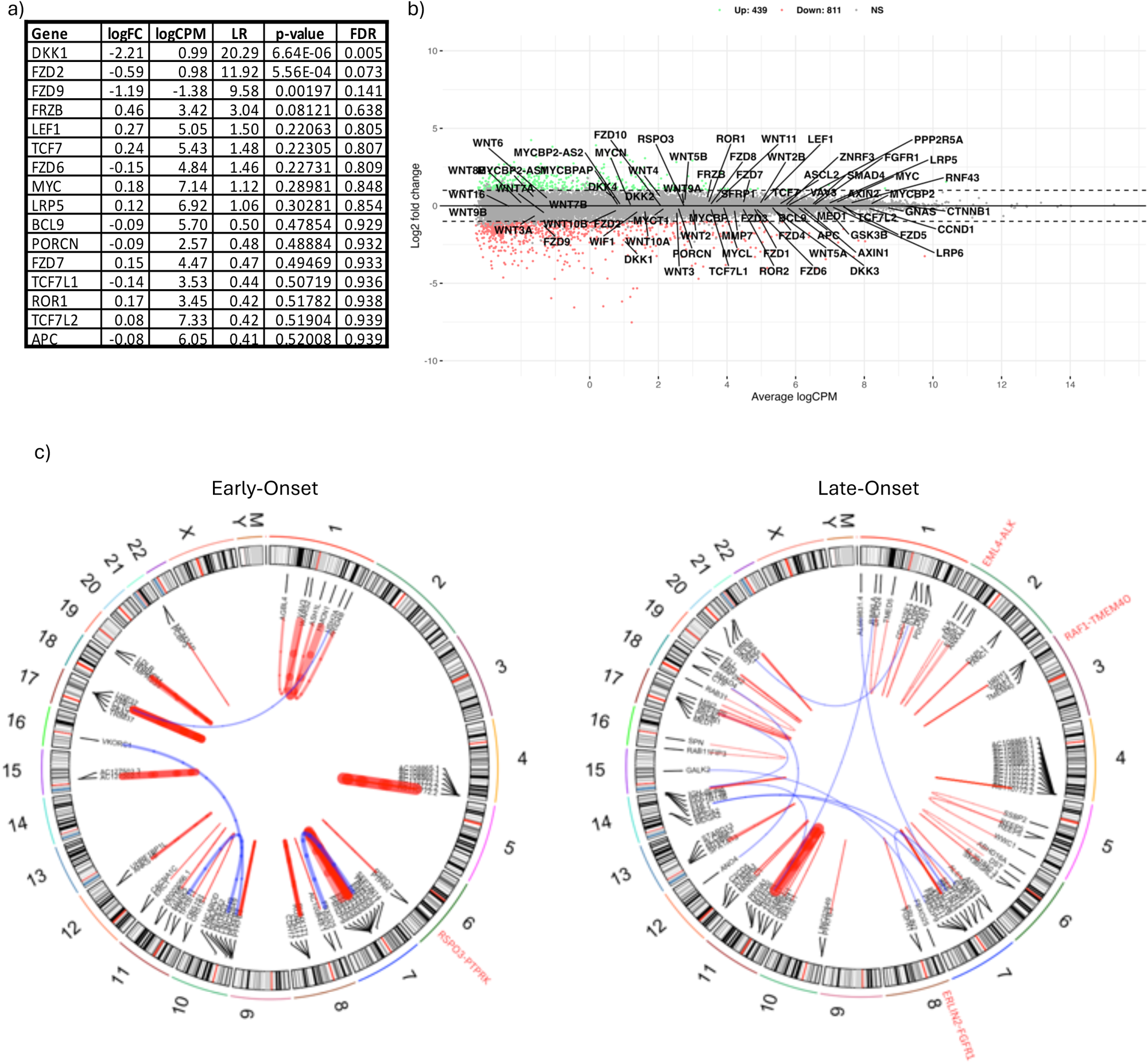
Differential gene expression (DGE) Analysis and Gene Fusion Analysis among CRC tumors from 30 Early-Onset and 37 Late-Onset Hispanic/Latino patients. **a)** Differentially expressed-related genes: This table shows the genes that are differentially expressed when comparing early-onset and late-onset CRC cases. **b)** Mean-Average (MA) plot: This scatter plot shows the upregulated and downregulated Wnt/β-catenin pathway genes according to the log2 fold change. **c)** These circle plots illustrate the gene fusions detected across 30 early-onset and 37 late-onset samples from our Hispanic/Latino (H/L) colorectal cancer (CRC) cohort. Clinically actionable gene fusions are highlighted in red, providing insights into potential therapeutic targets for this patient population.

### Transcriptomic comparisons with public databases: early-onset CRC tumors from Hispanics and Latino patients compared to Non-Hispanic White patients

When we compared early-onset CRC tumors in Hispanic/Latino patients to those in Non-Hispanic White patients, we observed distinct gene expression patterns in WNT pathway genes. Specifically, we performed differential gene expression analysis performed using RNA sequencing, (**Supplementary Fig. 3**). We analyzed 30 early-onset tumors from Hispanic/Latino patients and compared them to 30 early-onset tumors from Non-Hispanic White patients; data for the latter group was sourced from the TCGA-COAD and TCGA-READ projects. Tumors were matched based on key clinical parameters: tumor location, MSI status, age at onset, and TNM classification (**Supplementary Table 10**). According to our analysis, the two groups had significant differences in at least four WNT pathway genes—DKK2, DKK3, MYC, and LRP5; p-values were <0.0001. Additionally, we observed statistically significant differences in AXIN2, TCF7L2, APC, and ARID1A (p-values were <0.05) (**Supplementary Table 7**). Altogether, tumors in the Hispanic/Latino cohort appear to have unique functionality in the WNT pathway. Our results align with other studies where unique molecular characteristics appeared to contribute to documented disparities in CRC incidence and outcomes (Polakis et al. 2012; Siegel et al. 2022).

### Gene fusion analysis

Although gene fusions are relatively uncommon in CRC, the rise of fusion-targeted therapies highlights the importance of identifying clinically actionable fusions. To investigate these fusions, we conducted an analysis using STAR-Fusion in both early-onset and late-onset tumors (**Fig. 4c**). This comprehensive genomic coverage revealed 24 gene fusion events in early-onset tumors and 47 in late-onset tumors (**Supplementary Table 8**). Among these, 19 fusions were unique to the early-onset group, 41 were unique to the late-onset group, and 6 fusions were shared between the two groups. One particularly noteworthy fusion identified in the early-onset group is RSPO3. RSPO3 plays a direct role in the WNT signaling pathway—a critical pathway in CRC. The PTPRK-RSPO3 fusion has been well-documented to enhance WNT signaling and contribute to colorectal oncogenesis (Seshagiri et al., 2012; Kazanskaya et al., 2004). Additionally, the CDH17-RUNX1T1 fusion found in early-onset tumors involves RUNX1T1; this gene is associated with the transcriptional regulation of CTNNB1 (Clevers et al., 2012, Grinev et al 2021), which may indirectly influence WNT signaling. In contrast, the late-onset group did not exhibit any WNT pathway-related gene fusions. However, three notable non-WNT pathway fusions were identified, including two with known clinical relevance: EML4-ALK (Soda et al., 2007) and ERLIN2-FGFR1 (Wu et al., 2013). Additionally, the RAF1-TMEM40 fusion was identified, which may have potential clinical implications (Green et al., 2023; Yoshihara et al., 2014). These findings suggest distinct genetic drivers of CRC in the Hispanic Latino population. Because variations were observed depending on the age of onset, age-specific approaches are needed to better understand and treat CRC.

### WNT pathway alterations

One of our objectives was to understand how early-onset and late-onset colorectal cancer differed in terms of both the WNT pathway and MYC gene deregulation (**Fig. 5a**). Accordingly, we performed an integrated analysis of mutations, copy number variations, mRNA expression, and genetic similarity data. By categorizing samples according to the age of onset, we identified recurrent alterations in key WNT pathway genes: APC, TCF7L2, ARID1A, and SOX9 (**Fig. 5**).

**Figure 5:**
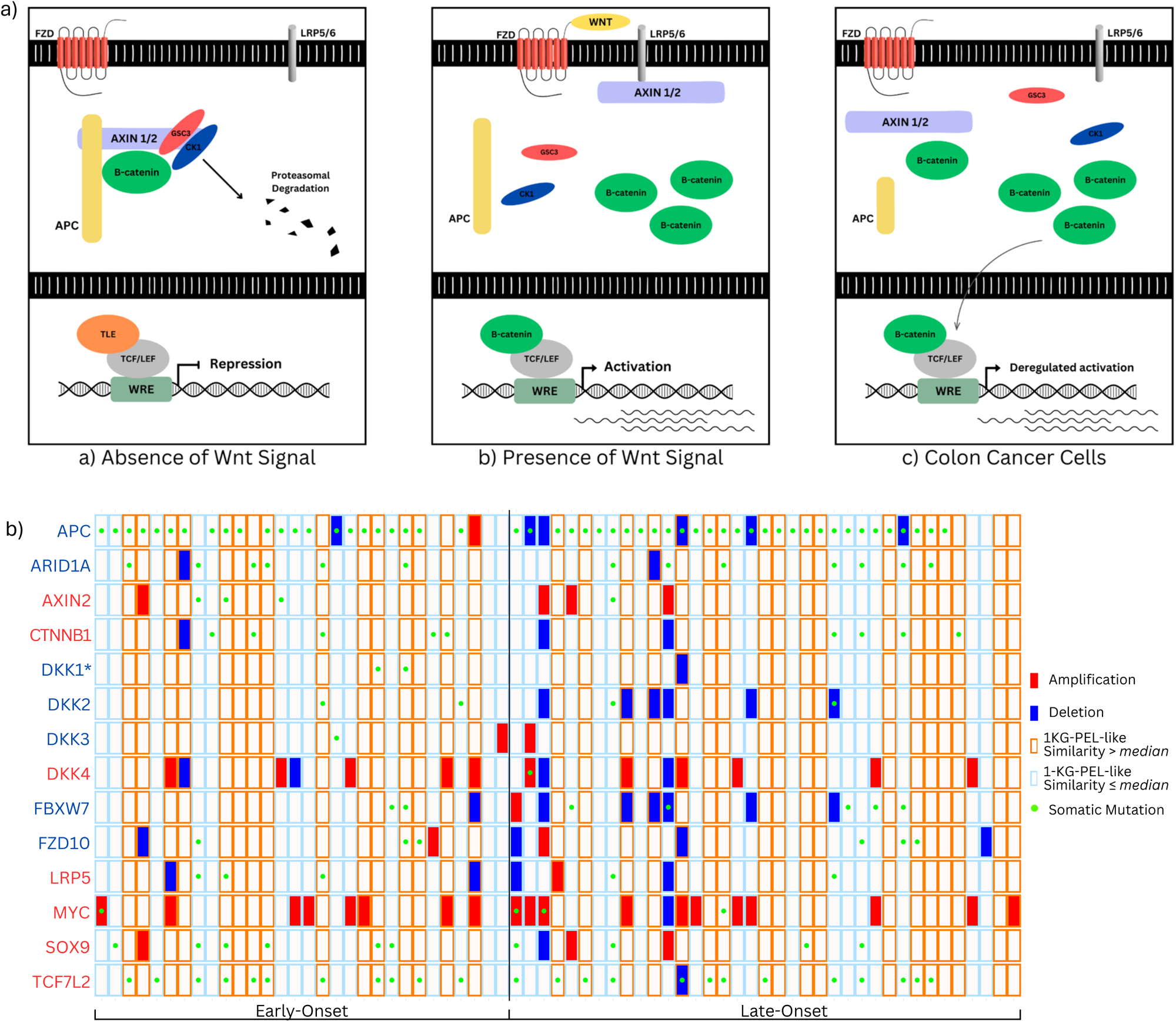
Integrative Genomic Alteration Patterns in Wnt/β-catenin Pathways Among CRC Tumors from 67 Hispanic/Latino Patients. **a)** The Wnt/β-catenin signaling pathway: **Panel A**: In the absence of Wnt signaling, β-catenin is degraded by a destruction complex. TCF/Lef proteins in the nucleus, along with the TLE co-repressor, suppress Wnt/β-catenin target genes. **Panel B**: Upon Wnt ligand binding to the FZD and LRP5/6 co-receptors, the destruction complex is inactivated, allowing β-catenin to accumulate and enter the nucleus. There, it binds TCF/Lef and recruits co-activators to activate target genes. **Panel C**: In colorectal cancers, APC truncations prevent β-catenin degradation, leading to its accumulation in the nucleus. This results in aberrant activation of Wnt/β-catenin target genes. **Abbreviations:** WRE: Wnt-responsive element; TCF/Lef: T-cell factor/Lymphoid enhancer factor; TLE: Transducin-like enhancer; FZD: Frizzled; LRP5/6: Lipoprotein receptor-related protein 5 or 6; AXIN1/2: Axis inhibition proteins 1 and/or 2; CRC: Colorectal cancer; APC: Adenomatous polyposis coli; GSK3: Glycogen synthase kinase 3; CK1: Casein kinase 1. **b)** This grid represents integrative genomic alteration patterns in the Wnt/β-catenin pathway among colorectal cancer (CRC) tumors from 67 Hispanic/Latino patients. Each column corresponds to an individual case, and each row represents a gene. The grid is divided into early-onset and late-onset. The color codes represent different attributes: red for amplifications, blue for deletions, an orange border for 1KG-PEL-like similarity proportion greater than the median, a light blue border for 1KG-PEL-like similarity proportion less than or equal to the median, and a bright green dot for somatic mutations. Genes are labeled in red for up-regulated and blue for down-regulated.

In our Hispanic/Latino cohort of CRC patients, both early-onset and late-onset tumors had a high prevalence of WNT signaling pathway perturbations: 96% of early-onset tumors and 97% of late-onset tumors (**Fig. 5b**). While deregulation of the WNT pathway is well-established in late-onset colorectal cancer (Marx et al., 2023), its role in early-onset tumors is poorly understood. Both cohorts displayed similar rates of 1KG-PEL-like: 50% of early-onset tumors and 49% of late-onset tumors had a 1KG-PEL-like proportion exceeding 55%. These results suggest that, despite the similarities in genetic similarity, there may be distinct biological mechanisms driving the development of colorectal cancer in early-onset versus late-onset cases within this population.

The early-onset cohort exhibited a higher proportion of APC mutations (59%) compared to SCNAs (7%). This trend was similarly observed in the late-onset cohort but with an even greater mutation rate of APC mutations (84%) and SCNAs (14%) (**Fig. 5b**). For the TCF7L2 gene, mutations were found in 37% of early-onset tumors without any SCNAs detected. The late-onset cohort showed a slightly higher TCF7L2 mutation rate (43%) and a small percentage of SCNAs (3%). ARID1A mutations occurred at comparable rates in both cohorts: 20% in early-onset with low SCNAs (5%); 19% in late-onset tumors also with low SCNAs (3%). Notably, SOX9 mutations were more prevalent in the early-onset group (20%) vs. late-onset group (13%). (SCNAs were more frequent in the late-onset group, 8.1% vs. 3%). These findings highlight distinct patterns of genetic alteration in WNT pathway when comparing early-onset to late-onset CRC.

Differences in DKK1 gene alterations may have significant implications for tumor behavior and progression. We observed higher DKK1 mutation rates in early-onset (7%) vs. late-onset tumors (0%). Thus, early-onset cases may experience unique disruptions in DKK1’s tumor-suppressive functions. Such disruptions can lead to unchecked WNT signaling and increased cell proliferation. On the other hand, late-onset tumors had 3% SCNAs that may also contribute to DKK1 dysregulation, albeit through a different mechanism. This is supported by RNAseq analysis: DKK1 expression exhibited downregulation, which may point to a loss of its inhibitory effect in the WNT pathway. Such downregulation may support tumor growth and progression in both early-onset and late-onset colorectal cancers. These findings underscore the pivotal role of DKK1 in regulating WNT signaling and highlight its potential influence on the distinct molecular characteristics observed in early-onset versus late-onset colorectal tumors.

## DISCUSSION

We comprehensively assessed both the MYC gene and WNT signaling pathway in 30 early-onset and 37 late-onset colorectal tumors (plus corresponding normal samples) from Hispanic/Latino individuals. Our analyses across the two cohorts uncovered important molecular differences. Notably, significant differences were observed between hypermutated and non-hypermutated tumors. For example, hypermutated tumors in the early-onset cohort were predominantly located in the right colon; they had higher levels of 1KG-PEL-like vs. non-hypermutated counterparts. In contrast, non-hypermutated tumors in the early-onset cohort were mainly found in the left colon; they had lower levels of 1KG-PEL-like vs. hypermutated counterparts. We do not know why hypermutated early-onset tumors are mainly the right colon and exhibit high 1KG-PEL-like. We will aim future studies at understanding these differences at a molecular level to identify potential pathways for targeted interventions. However, these results provide evidence that tumor characteristics are significantly influenced by both genetic and ethnic backgrounds.

As we continued to compare early-onset and late-onset CRC, we found that late-onset patients were more likely to present with a higher tumor mutational burden (TMB). Specifically, TMB was approximately four times more frequent in late-onset patients vs. early-onset patients. Thus, early-onset patients who have low TMB may exhibit distinct tumorigenic pathways or genetic mechanisms. Such differences could influence responses to immunotherapy – e.g., lower TMB is often associated with poorer treatment outcomes. However, the precise magnitude of this risk increase is uncertain due to the broad confidence interval observed in the data. However, others (Marx et al., 2023) have observed how early-onset CRC (vs. late) appears to exhibit distinct biological characteristics. Understanding the unique mutational profiles of early-onset CRC thus will be key when tailoring therapies for this subgroup.

We compared mutational frequencies of MYC and WNT pathway genes in our early-onset Hispanic/Latino cohort with those in six publicly available CRC genomic datasets: MSK-NatCommun, MSK-JNCI, DFCI-CellReports, TCGA-PanCancerAtlas, MSK-Gastroenterology, and MSK-JCO-PrecisOncol (Table 2 and Supplementary Fig. 2). These databases predominantly represent non-Hispanic White patients. Our findings revealed significant differences between our cohort and the publicly available early-onset databases in the mutational frequencies of 13 WNT pathway genes. To further investigate these differences, we also used the AACR Project GENIE dataset; this database is stratified by self-reported race and predominantly represents non-Hispanic White patients. Our analysis uncovered significant differences in key WNT pathway genes: DKK1, DKK2, FZD10, and LRP5. Although there were notable similarities in mutation frequencies across these large datasets, our analysis also highlighted distinct molecular characteristics unique to the early-onset Hispanic/Latino cohort. These findings emphasize the need to more comprehensively include diverse early-onset populations in genetic research. Such inclusion is essential for deepening our understanding of cancer disparities and identifying potential targets for precision medicine in CRC.

To further probe the roles of both the MYC gene and WNT signaling pathway in CRC subpopulations, we compared early-onset and late-onset Hispanic/Latino patients by combining whole-exome sequencing with integrative genomic analysis. Notably, we observed mutations and somatic copy number alterations (SCNAs) in both MYC and the WNT pathway for 97% of tumors in the early-onset cohort. Such genetic factors may be critical in the development of CRC within this demographic. Moreover, in our transcriptomic analysis of early-onset CRC (vs. late-), we observed marked deregulation of the DKK2 gene. DKK2 not only showed the highest proportion of 1KG-PEL-like at 78.2%, but it was also associated with the second-lowest average age among early-onset patients (**Supplementary Table 6**). Given the statistical significance of this finding, DKK2 may be critical for tumorigenesis of early-onset CRC – particularly in populations with a high degree of 1KG-PEL-like. DKK2 functions as a tumor suppressor within the WNT pathway; its deregulation may indicate a distinct molecular mechanism driving cancer development in younger individuals modulated by both genetic and genetic similarity factors. Our findings highlight the necessity of integrating genetic and demographic variables to fully understand the unique characteristics and risks associated with early-onset CRC, especially in diverse populations.

Our early-onset Hispanic/Latino cohort (vs. non-Hispanic White) displayed unique patterns of gene deregulation within the WNT pathway. Specifically, we observed alterations in the following tumor suppressor genes: DKK2, DKK3, AXIN2, APC, and ARID1A. Alongside, we also observed alterations in oncogenes like LRP5, TCF7L2, and MYC. We now briefly describe the roles these genes play in the WNT pathway and tumor development. The tumor suppressor genes DKK2 and DKK3 are known to inhibit the WNT signaling pathway (Mao et al. 2002), which is frequently dysregulated in CRC. When these genes are inactivated, the resulting dysregulation can lead to unchecked cell proliferation. Similarly, ARID1A (Jones et al. 2010), another tumor suppressor involved in chromatin remodeling, was frequently mutated in our cohort. This finding aligns with its known role in disrupting gene expression and promoting tumor progression in CRC. Interestingly, both ARID1A and the well-established oncogene MYC (Dang et at. 2012) were significantly deregulated in early-onset tumors. This observation is consistent with previous reports that ARID1A suppresses MYC transcription in CRC (Nagl et al. 2006); it may be one mechanism underlying EOCRC for Hispanic/Latino patients. Moreover, LRP5 functions as a co-receptor in the WNT pathway and can act as an oncogene depending on the cellular context (Katoh et al. 2017). The tumor suppressors AXIN2 (Jho et al. 2000) and APC (Groden et al. 1991) play critical roles in regulating β-catenin degradation, thereby inhibiting WNT signaling. Mutations in these genes lead to the accumulation of β-catenin and the activation of WNT target genes, such as MYC, which drives CRC development. Finally, TCF7L2, generally considered an oncogene, partners with β-catenin to activate the transcription of WNT target genes, further contributing to tumorigenesis (Schwitalla et al. 2013). Overall, our findings highlight the complexity and diversity of WNT pathway alterations in CRC, particularly in early-onset cases among Hispanic/Latino populations. These results underscore the need for further research to better understand the ethnic and age-related differences in CRC pathogenesis.

To the best of our knowledge, this is the first report to reveal molecular differences between early-onset and late-onset CRC in Hispanic/Latino patients. These differences are particularly significant given that both early-onset and late-onset CRC tumors are currently treated with the similar standard-of-care. For example, early-onset tumors in our cohort (vs. late) exhibited distinct patterns of copy number variations and gene expression profiles. All early-onset tumors showed perturbations in one or more genes within the WNT signaling pathway; except for one sample with low 1KG-PEL-like, most of these early onset samples had high 1KG-PEL-like. From a therapeutic perspective, our data also highlight a clinically actionable gene fusion, RSPO3-PTPRK (Storm et al. 2016), identified in an early-onset sample. This fusion typically leads to the overactivation of the WNT signaling pathway, which plays a critical role in colorectal tumorigenesis. The presence of this fusion suggests that tumors harboring it may be particularly sensitive to therapies targeting the WNT pathway. RSPO3 fusions have been associated with a distinct subset of colorectal cancers that could benefit from targeted therapies, such as porcupine inhibitors, which inhibit WNT ligand secretion, or other WNT pathway inhibitors. Although RSPO3-PTPRK fusion was found in one sample, it still highlights how targeted therapies may be used in the future to inhibit the WNT pathway in EOCRC in the Hispanic/Latino population.

## CONCLUSION

This study provides critical insights into the molecular characteristics of early-onset CRC in Hispanic/Latino populations. We compare this subgroup to late-onset CRC in the same demographic and early-onset CRC in Non-Hispanic White patients. We also show how integrating multi-omics analysis with genetic 1KG-PEL-likeproportions can be a valuable resource for understanding potential therapeutic targets in an underrepresented minority group. To the best of our knowledge, this is the first integrative report to both map the multi-omics landscape and probe associations between genetic 1KG-PEL-likeand EOCRC in Hispanic/Latino populations. Our findings highlight significant biological differences that may impact screening and treatment strategies. These differences underscore the importance of precision medicine approaches tailored to the unique genetic and molecular profiles of this population. Further research with larger sample sizes is essential to fully elucidate the genomic heterogeneity and the role of 1KG-PEL-likein the development of early-onset CRC tumors in Hispanic/Latino populations. Such efforts can help reduce cancer disparities by ensuring that advances in cancer prevention, diagnosis, and treatment are both accessible and effective for historically underserved groups. Ultimately, our goal is to improve clinical outcomes through more personalized therapeutic interventions, thereby contributing to the reduction of health disparities in cancer care.

## MATERIALS AND METHODS

### Study population

Participants for this study were recruited through the ENLACE study, conducted under the Center for Optimizing Engagement of Hispanic Colorectal Cancer Patients in Cancer Genomic Characterization Studies (COPECC). Recruitment took place at the oncology clinics of the University of Southern California (USC) Norris Comprehensive Cancer Center and the Los Angeles General Medical Center. Eligible participants were individuals aged 18 years or older who self-identified as Hispanic/Latino and had a confirmed diagnosis of colorectal cancer. The USC COPECC center (https://usccopecc.org) is a member of the Participant Engagement and Cancer Genome Sequencing (PE-CGS) Network (https://pe-cgs.org), which is part of the Cancer Moonshot Initiative^SM^ (https://www.cancer.gov/research/key-initiatives/moonshot-cancer-initiative) aimed at accelerating cancer research progress. For this study, a total of 67 patients diagnosed between 2022 and 2024 were included, consisting of 30 early-onset colorectal cancer (EOCRC) patients— defined as those diagnosed before age 50—and 37 late-onset patients. This study was approved by the Institutional Review Board of the University of Southern California.

### Data collection

All participants completed questionnaires, donated blood samples, and provided access to their tumor tissue through the USC COPECC Patient Engagement Unit (https://usccopecc.org/patient-engagement-unit/). This initiative aims to develop a patient engagement framework specifically tailored to meet the unique needs of Hispanic/Latino cancer patients.

### Clinical data

Clinical data was obtained for all patients through medical chart abstraction. Specifically, data on tumor location, tumor stage (TNM stage), MSI, primary site of diagnosis, colon side, pathological group stage at diagnosis, and medications used were collected.

### DNA and RNA extraction and quality control

Tumor tissue and blood samples were collected from study participants according to standard operating procedures through the USC COPECC center Genome Characterization Unit (https://usccopecc.org/genome-characterization-unit/).

### DNA Isolation

Genomic DNA and RNA was extracted from tissue using the Covaris truXTRAC© total NA Column kit (Woburn, MA) based on the manufacturer’s recommendations. Briefly, tissue samples were placed into Covaris Adaptive Focused Acoustics (AFA) and suspended in Lysis buffer followed by Proteinase K digestion. Tissue mixtures were then equally split for separate DNA and RNA extraction. Tissue mixtures were then ultrasonically emulsed using the Covaris E220 System. DNA or RNA sample emulsions were then separately chemically de-crosslinked and pipetted onto appropriate spin columns. DNA and RNA were then collected using provided elution buffers. DNA and RNA were quality assessed and quantified using both the (Thermo Fisher Scientific, Inc. Waltham, MA) and the Genomic DNA Screen Tape Assay utilizing the Agilent 4200 TapeStation System (Santa Clara, CA). DNA and RNA samples were stored at −80°C.

### Whole exome and RNA sequencing

For Whole exome sequencing we utilized a custom expanded exome bait set (Agilent Technologies, Inc.). Briefly, components of the expanded exome included the following probe groups: original baits from SureSelect Human All Exon V6, (Agilent Technologies, Inc.) and custom baits for select genomic regions.(Manojlovic et al. 2017) Genomic DNA in the amount of 50-200 nanograms (ng) from colorectal tumor tissue and paired uninvolved (normal) colorectal tissue from each case was sheared in 50 microliters (ml) of TE low EDTA buffer employing the Covaris E220 system (Covaris, Inc., Woburn, MA) to target fragment sizes of 150 – 200 bp. Fragmented DNA was then converted to an adapter-ligated whole genome library using the Kapa Hyper Prep Library Prep kit (Kapa Biosciences, Inc., Wilmington, MA) according to the manufacturer’s protocol. SureSelect XT Adaptor Oligo Mix was utilized in the ligation step (Agilent Technologies, Inc.). Individual tumor adapter-ligated libraries were enriched into the exome capture reaction, and for germline each adapter-ligated library was pooled before proceeding to capture using Agilent’s SureSelect Human All Exon V6 + custom probes capture library kit. Samples that had successful libraries created were then sequenced on Illumina MiSeq technology for quality control to assess the ability of the libraries to be sequenced. Subsequently, each library was pooled and sequenced on Illumina’s NovaSeq 6000 (Illumina, San Diego, CA) using 300 cycle kit. Raw FASTQs were generated using the industry standard BCL2FASTQ v1.8.4. Mean target coverage was 105X for tumor samples and 55X for uninvolved control samples.

RNA was extracted from cells from colorectal cancer tissues samples and analyzed by RNA-seq. Sequencing libraries prepared with the TruSeq Stranded Total RNA kit (Illumina Inc), from 1 μg total RNA.

### Whole exome sequencing analysis

All sequencing reads were converted to industry standard FASTQ files using the Bcl Conversion and Demultiplexing tool (Illumina, Inc). Sequencing reads were aligned to the GRCh38 reference genome using the MEM module of BWA v0.7.17 (Li et al. 2009) and SAMTOOLS v1.9 [117] to produce BAM files. After alignment, the base quality scores were recalibrated and joint indel realignment was performed on the BAM files using GATK v4.0.10.1. (McKenna et a. 2010) Duplicate read pairs were marked using PICARD v2.18.22. (Broad Institute et al 2010) Final BAM files were then used to identify germline and somatic events. Germline SNP and INDELS were identified using GATK haplotype caller in the constitutional sample.

### Gene mutations analysis

Somatic variant callers identified single nucleotide variants (SNVs) and small insertions and deletions (indels). Somatic mutation analysis was performed by identifying overlapping variant calls generated by Strelka(Kim et al. 2018) and Mutect.(Benjamin et al. 2019) Due to the diffuse nature of these tumor samples, we retained somatic mutations that had a minor allele frequency of 4%, and manually reviewed variants in the KRAS region using IGV.(Robinson et al. 2017) We compared these mutations against germline results to confirm and verify the somatic mutations. After filtering and manually reviewing somatic variants, VCFs were then annotated with Ensembl’s VEP (McLaren et al 2016) tool and converted to the MAF file format for visualization purposes. MAFtools (Mayakonda et al. 2018) was utilized in the generation of oncoplots, annotated with demographic information associated with each sample.

### Somatic Copy-Number Alterations (SCNAs) analysis

For chromosomal and sub-chromosomal changes analysis we used GISTIC2.0 (Mermel et al. 2011) best practices to identify genes targeted by somatic copy-number alterations (SCNAs) and SCNA profiles into underlying arm-level and focal alterations.

### Genetic similarity analysis

We used GATK HaplotypeCaller v4.0.10.1 to generate germline VCF files for genetic similarity analysis. Known population genotype data were downloaded from the 1000 Genomes Project phase 3 and grouped by super population for genetic similarity analysis.(1000 Genomes Project Consortium et al. 2015) VCF subsetting and merging were performed using VCFtools v0.1.17 and SnpSift v4.3t.(Danecek et al 2011, Cingolani et al 2012) VCF genotype allele coding (0/1) was converted to numeric (012) using VCFtools and PLINK v1.90b6.7.(Chang et al 2015) All genetic similarity analyses were performed on autosomal chromosomes. Global population admixture was estimated using STRUCTURE v2.3.4.(Falush et al. 2003) We subset and merged all VCF files by a list of 1766 genetic similarity-informative markers (AIMs). The result was converted to Structure format by PLINK. To run STRUCTURE, we set up population numbers k=5, NUMREPS=2000, and BURNIN=50000. Principal component analysis (PCA) was performed on the same dataset by R function prcomp. Local genetic similarity analysis was performed using the tool LAMP-LD v1.3.(Baran et al. 2012) Five super population genetic similarity haplotype files were prepared using an expanded AIM list of 20803 SNPs (single nucleotide polymorphisms). To run LAMP-LD, we set the window length to 50 SNPs, and the number of states was set to 20. All genetic similarity analysis results were visualized using R v3.6.0 ggplot2 v3.4.1(Wickham et al. 2016) package. For initial analysis, genotypes for a total of 1,766 AIMs were extracted and used for estimating global genetic genetic similarity based upon five genetic similarity super continental populations including 1000 Genomes Project Peruvian-in-Lima-like (1KG-PEL-like), 1000 Genomes Project European-like, (1KG-EUR-like), 1000 Genomes Project African-like (1KG-AFR-like), 1000 Genomes Project South-Asian-like (1KG-SAS-like), 1000 Genomes Project East-Asian-like (1KG-EAS-like) using STRUCTURE (**Fig. 1**). We further used LAMP-LD to estimate local genetic similarity for 20,803 SNPs extracted from WES data across the genome for deducing chromosome level admixture for each individual relative to the same five super populations (**Supplemental Fig. 1**).

### RNA sequencing analysis

Alignment of raw sequence reads to the human transcriptome (hg38) was performed via Rsubread (Liao et al. 2019) and transcript abundance estimates were normalized and differentially expressed genes (DEG) identified using a standard edgeR pipeline. Functional annotation of gene sets: Pathway enrichment analysis and gene set enrichment analysis (GSEA) were performed using gene sets from the Molecular signatures database (MSigDB). In addition, R package gage (Generally Applicable Gene-set Enrichment) where used to identify enriched pathway and pathview to visualize these pathways. For transcript-aware analyses, the FASTQ files were aligned with salmon (Patro et al 2017) and differentially enriched transcripts were identified using DRIMSeq (Nowicka et al. 2016) in a similar workflow to edgeR (Long et al. 2021).

### Gene fusions analysis

For gene fusion we used STAR-Fusion that is built on top of the STAR aligner according with its best practices. (Dobin et al. 2013) We ran STAR version 2.5.3a, STAR index was created, and reads were aligned to the same human reference genome than used for the WES analysis.

### Statistical analysis

In this study, we included 67 subjects recruited as part of the ENLACE study from USC COPECC, of which all passed the genomic sequencing quality controls (**Supplemental Table 1**). Clinical and demographic data were evaluated from each participant. The demographic variables evaluated in the study included gender, age, race and ethnicity. In addition, the following clinical data, including tumor characteristics were evaluated: tumor location, tumor stage (TNM stage), MSI, primary site of diagnosis, colon side, pathological group stage at diagnosis, mutation status of CRC related genes and medications used. Genetic similarity was modeled as a categorical variable (dichotomous for each genetic similarity population: less or equal than median genetic similarity levels). Demographic, clinical and genomic characteristics among CRC cases were evaluated according to CRC status using Pearson’s chi-square or Fisher’s exact test, as appropriate for categorical variables. Overall, since a previous efforts show no difference between unadjusted and adjusted ORs, we present unadjusted ORs and its 95% confidence interval (CI) for the association of genetic similarity and each tumor characteristic.

### WES Comparison with Public Databases

cBioPortal (Cerami et al. 2012) for Cancer Genomics that is highly regarded resource for interactive exploration of multidimensional cancer genomics datasets was used to extract genomic data from publicly available datasets and comparisons were evaluated using Pearson’s chi-square or Fisher’s exact test, as appropriate.

### Transcriptomics Comparison with Public Databases

Transcriptomic data of 67 Non-Hispanic White was extracted from public databases projects TCGA-COAD and TCGA-READ projects. All individual demographic and clinical data from the 67 CRC tumors from Non-Hispanic White were matched to the same data from our 67 Hispanic/Latino CRC tumor samples. Differential Gene expression analyses were performed as mentioned before in the RNA sequencing analysis section.

## Data Availability

All data produced in the present study are available upon reasonable request to the authors

## Abbreviations

CRC: Colorectal Cancer
Hispanic/Latino: 
Non-Hispanic White: Non-Hispanic White
MSI: Microsatellite instability
DGE: Differential Gene Expression
SCNA: Somatic Copy Number Alteration
SNP: Single nucleotide polymorphism
US: United States

## Acknowledgements

The authors would like to thank the Department of Integrative Translational Sciences at City of Hope (COH), the Cancer Control and Population Sciences program (P30CA033572) within the City of Hope Comprehensive Cancer Center, as well as, the NIH NCI grant project number U2CCA252971, the genomic characterization unit and the center for Optimizing Engagement of Hispanic Colorectal Cancer Patients in Cancer Genomic Characterization (COPECC) Studies in the University of Southern California, Los Angeles. In addition, the authors would like to thank the patients, families, advocates, recruiters and all the personnel involved in the study for their contribution in this study.

